# Parkinson’s disease psychosis associated with accelerated multi-domain cognitive decline

**DOI:** 10.1101/2024.02.16.24302938

**Authors:** Sara Pisani, Luca Gosse, Dag Aarsland, K. Ray Chaudhuri, Clive Ballard, Dominic ffytche, Latha Velayudhan, Sagnik Bhattacharyya

## Abstract

Cognitive deficits are associated with poor quality of life and increased risk of development of dementia in people with Parkinson’s Disease (PD) with psychosis. However, the pattern of progression of cognitive decline within PD psychosis remains unclear. Here, we examined this using data from the Parkinson’s Progression Markers Initiative study.

We obtained data on drug-naïve PD patients (n=676) and healthy controls (HC, n=187) who underwent baseline and follow-up (year 1 to 5) assessments. We classified PD patients into PD without psychosis (PDnP) and PD with psychosis (PDP) using the MDS-UPDRS part I hallucinations/psychosis item. We examined all cognitive measures assessed at each time point. We used linear mixed-effect models with restricted maximum likelihood. We examined the role of age, sex, ethnicity, education, and neuropsychiatric and PD-specific symptoms as covariates of interest.

There were no baseline differences on any cognitive measures between PD patient groups. There were differences in cognitive performance between PD and HC across the majority of the assessments. PDP patients showed a more prominent cognitive decline from baseline to year 5 compared with PDnP across most domains even after controlling for socio-demographics, depression, sleepiness, REM behaviour sleep disorder, and motor symptom severity (immediate recall, b=-0.288, *P=*0.003; delayed recall, b=-0.146, *P=*0.003; global cognition, MoCA, b=-0.206, *P <*0.001; visuo-spatial, b=-0.178, *P=*0.012; semantic fluency, b=-0.704, *P=*0.002; processing speed, b=-0.337, *P=*0.029).

Over five years, PD psychosis patients showed worsening of performance in semantic aspects of language, processing speed, global cognition, visuo-spatial abilities, and memory sub-components, regardless of socio-demographic characteristics, neuropsychiatric and motor symptoms. Collectively, these cognitive domains, particularly semantic aspects of language may therefore play an important role in PD psychosis and warrant further investigation.

## Introduction

Psychotic symptoms such as hallucinations and delusions are some of the most common and debilitating non-motor symptoms in patients with Parkinson’s Disease (PD).^1,2^ People with PD psychosis (PDP) have poor quality of life and are at increased risk of hospitalisation and dementia.^3,4^ PDP is associated with cognitive deficits such as impairments in attention, global cognition, executive functions, and processing speed, which may further lead to deterioration in quality of life and increased risk of dementia.^5,6^

It is clinically recognised that people with PD psychosis report worse cognitive performance compared with PD patients who do not have such symptoms ^7–9^ consistent with two recent meta-analyses.^10,11^ Pooling data from cross-sectional comparisons, they found that functioning across all major cognitive domains, i.e., attention, global cognition, memory, perception, and executive functions, is impaired in PDP compared to PD patients without psychosis patients, with executive function and attention being the most affected domains.^10,11^ They also reported that age at testing, ^10,11^ depression and PD duration ^11^ had a moderating effect on some of these deficits. Specifically, episodic memory was the most likely impaired domain in PDP as shown by a sub-group analysis conducted in the same samples of patients with a PD duration of 6-9 years. Encoding and retrieval, domains of episodic memory,^12^ phonemic and semantic fluency, perception (namely dorsal and ventral stream associated visuo-spatial abilities, and low level visual apperception), construction (namely copying) were also impaired in PDP compared to PD patients without psychosis, suggesting a potential role of these domains in the symptomatology of psychosis in PDP. However, inference on the progression of cognitive functioning over time is challenging on the basis of evidence from cross-sectional studies. Among the handful of longitudinal studies in this field, the PRIAMO study ^13^ showed that PD patients who developed psychosis symptoms (n=37) had worse performance in global cognition (as indexed using the Mini Mental State Examination (MMSE)) at 12 months and 24 months follow-up. Similarly, in their clinic-based retrospective sample, Muller et al.^14^ found that PD patients who had VH (n=18) at follow-up (mean follow-up duration: 2.8 years) had worse performance at baseline and follow-up compared with PD patients without psychosis (n=15; mean follow-up duration: 4.7 years) on the measures of processing speed and more broadly on executive functions. Goetz et al. ^15^ also examined differences between PD patients with and without hallucinations over a period of 6 years. 37 out of 60 PD patients developed hallucinations across different sensory modalities (e.g., visual vs non-visual), however these two groups did not differ on cognition as measured with the MMSE after 6 years from baseline. Using data from the Parkinson’s Progression Markers Initiative (PPMI) study^16^ (n=131 PD patients), a 5-year longitudinal study in patients with de-novo PD, another study found that a higher proportion of those who developed psychotic symptoms reported subjective cognitive decline compared to those who did not, without any significant group difference in cognitive task performance at follow-up.^17^ Using the same PPMI study, ffytche et al.^18^ showed that 115 PD patients who developed minor illusions (onset at 19.5 months follow-up) differed on neuropsychiatric symptoms, and olfaction at baseline compared with PD patients who never developed such symptoms. Although there was no difference in the slope of cognitive decline prior to developing psychosis symptoms between groups, those who developed more severe symptoms of psychosis (n=21, 4.9%) also showed worse performance on the Benton Judgement of Line Orientation (BJLOT) compared with PD patients, at baseline.

While it is generally accepted that there is a progressive decline in cognitive function in people with PD and that this may be greater in PD patients with psychosis,^19–21^ to the best of our knowledge the longitudinal course of cognition has not been systematically examined before. Therefore, to address this gap in evidence here we compare the longitudinal course of cognitive task performance across a range of measures over the 5 years of follow-up from cohort inception in people recruited into the PPMI study. Based on previous literature, we predicted that cognitive task performance will show a greater decline in PD patients compared to healthy individuals. Our main hypothesis of interest was that this decline will be even greater in PD patients who later on develop psychosis compared to those who do not develop psychosis over the follow-up period, even after controlling for potential confounders.

## Materials and methods

### Participants

The PPMI study enrolled newly diagnosed unmedicated patients with Parkinson’s Disease and age- and gender-matched healthy controls. Details of eligibility criteria, objectives and methodology have been published elsewhere^16^ and can be also found on www.ppmi-info.org/study-design (ClinicalTrials.gov NCT01141023). In brief, PD patients included were drug-naïve, within 2 years of PD diagnosis, with a Hoen & Yahr stage <3, 30 years of age or older, had either at least two PD symptoms (e.g., slowness of movement, tremor, or rigidity), or single asymmetric resting tremor or slowness of movement. Healthy controls were included if they were 30 years of age or older, with no evidence of neurological disorder or a first-degree relative with PD. PD patients were excluded if they had a diagnosis of dementia and healthy controls were excluded if they had a Montreal Cognitive Assessment (MoCA) score ≤ 26 upon study enrolment. For the purpose of this study, we excluded <u>s</u>ubjects <u>w</u>ithout <u>e</u>vidence of <u>d</u>opaminergic <u>deficits</u> (i.e., SWEDD) because of their prognostic and clinical differences from those with an idiopathic PD diagnosis.^22–24^ We also excluded individuals who transitioned to dementia within the 5 study years as we wanted to examine the hypothesised differential decline between PD psychosis and PD without psychosis patients, without it being potentially confounded by participants being in the early stages of dementia. These data were accessed and downloaded on 1^st^ February 2023 (Figure 1). The PPMI study was approved by the institutional review boards at each study site contributing data and participants provided written informed consent.^16,25^ The data used in this analysis are openly available from the PPMI study.

**Figure 1.**
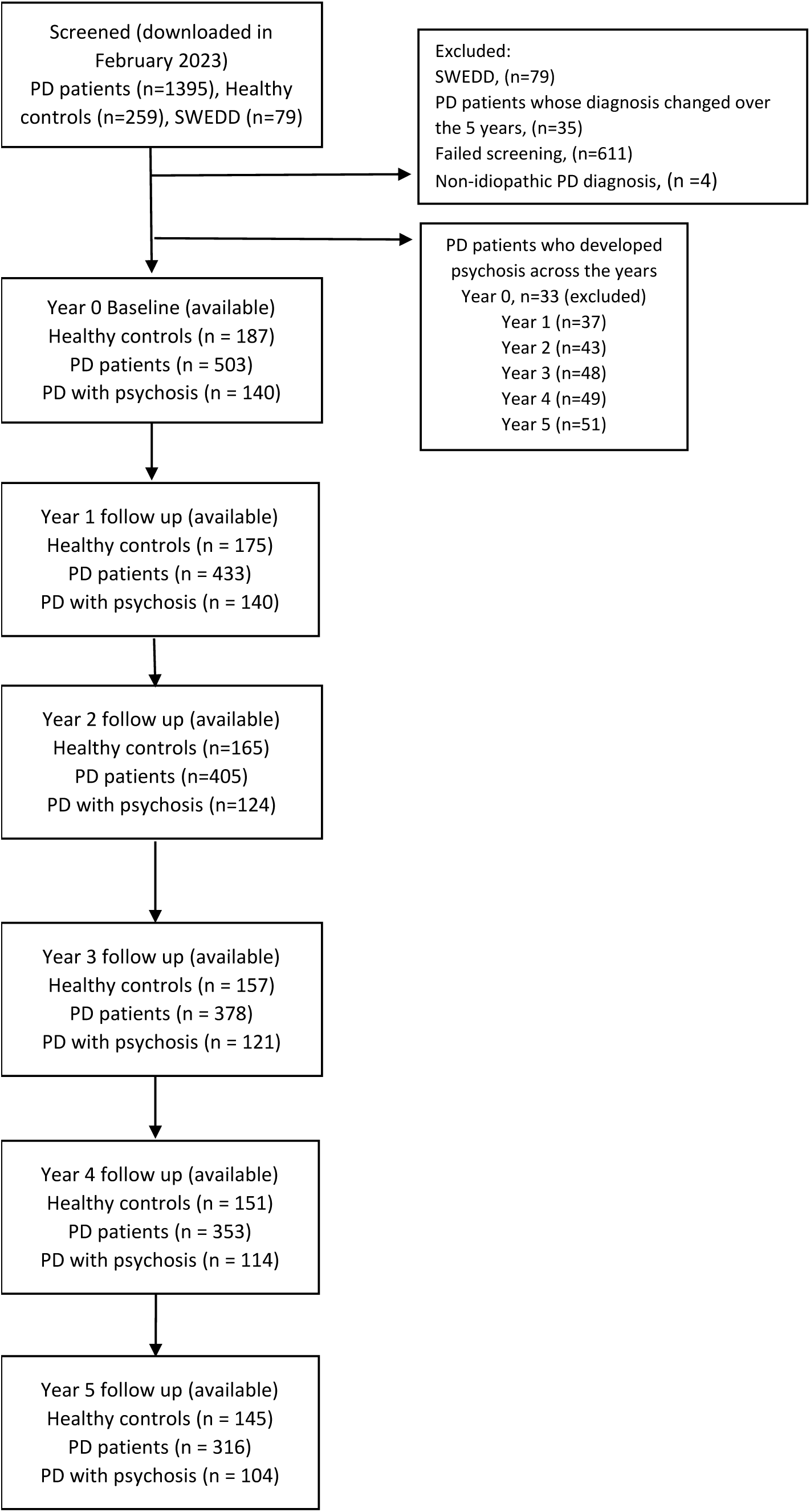
Participant flow chart. Chart for flow of participants enrolled in the PPMI study.

### PD-specific outcome measures

PD severity was assessed using the Movement Disorders Society Unified Parkinson’s Disease Rating Scale (MDS-UPDRS) and Hoen & Yahr stages.^26^ Assessments of tremor and rigidity were also included across study visits. PD medication use was assessed by converting to a composite measure of Levodopa equivalent daily dose (LEDD). Information about LEDD was extracted from the most recently released dataset and followed the recommendation of the PPMI study group (see Supplementary Material 1 for more information, and for group differences on LEDD). Autonomic symptoms were measured with the Scale for Outcome in Parkinson’s Disease – Autonomic (SCOPA-AUT).^27^

### Classification of PD psychosis patients

PD patients were classified into PD psychosis (PDP) and PD without psychosis (PDnP) based on previous work ^18^ using the MDS-UPDRS part I hallucinations/psychosis item, which measures the presence of visual hallucinations and paranoid thoughts. On this item, patients may receive a score out of 5: 0=normal, no hallucinations or psychotic behaviour, 1=slight, illusions or non-formed hallucinations, but patient recognises them without loss of insight, 2=mild, formed hallucinations independent of environmental stimuli, no loss of insight, 3=moderate, formed hallucinations with loss of insight, 4=severe, patient has delusions or paranoia. If PD patients reported a score ≥1 at any of the study visits, they were considered as PDP (i.e., with psychosis). We have excluded PD patients who reported psychosis symptoms at baseline (n=33) because the purpose of this study was to examine the decline in PD patients with and without psychosis with both groups starting off on a similar level. For more information on this patient group, please refer to Supplementary Material 2. Three groups were therefore created:

‣ *HC:* healthy controls (no PD symptoms)
‣ *PDnP:* PD patients with a score of 0, i.e., without reports of psychotic symptoms throughout the study
‣ *PDP:* PD patients with a score ≥1, i.e., with presence of psychosis symptoms which included hallucinations and delusions, at any time point throughout the study.

### Cognitive and neuropsychiatric assessments

Global cognition was assessed using the MoCA.^28^ Episodic memory was assessed with the Hopkins Verbal Learning Test – Revised (HVLT-R),^29^ which measures memory sub-domains such as immediate and delayed recall, recognition and discrimination. The PPMI study also included other assessments such as Symbol Digit Modality (SDM) ^30,31^ which measures processing speed, Letter Number Sequence (LNS) ^32^ for working memory (i.e., manipulation) and Semantic Fluency tests (i.e., category based) ^33^ to assess semantic aspects of language, and Benton Judgement of Line Orientation (BJLOT) ^34^ which is a measure of visuo-spatial ability. A range of neuropsychiatric symptoms were included as covariates in analyses: depressive symptoms indexed using the Geriatric Depression Scale 15 items (GDS-15)^35^; anxiety traits and states measured using the State-Trait Anxiety Inventory (STAI)^36^; and sleep indexed using the Epworth Sleepiness Scale and a REM sleep behaviour disorder (RBD) questionnaire.^37,38^

### Statistical analysis

Baseline characteristics were analysed using Analysis of Variance (ANOVA) or its non-parametric equivalent (e.g., Kruskal-Wallis χ^2^) as appropriate. We report baseline characteristics for all major cognitive assessments, neuropsychiatric measures, and PD-related assessments. We examined all study visits from baseline to final follow up at year 5. We examined the HVLT-R, Symbol Digit Modality (SDM), semantic fluency test, Letter Number Sequence (LNS), Benton Judgement of Line Orientation (BJLOT) and MoCA as outcome measures to assess cognitive performance over 5 years and the difference in such performance between PDP and PDnP patients. The PPMI study include age- and sex-matched healthy controls which were included in the present analysis. We employed linear mixed-effect analyses with restricted maximum likelihood (REML) using the *lmerTest* package ^39^ in R (version 4.0.3).^40^ In all analyses, we entered group as a between-subject factor (i.e., HC, PDnP, and PDP) and year (i.e., treated as continuous variable, baseline as year 0 to year 5) as a within-subject factor, and patient number as a random effect variable. Significance (a two-sided alpha level of 0.05) was estimated using Satterthwaite’s method. For all cognitive tasks, we first tested a simple (unadjusted) model and then included confounders of interest in a separate linear model. We included sociodemographic (i.e., age, sex, ethnicity, and years of education) as well clinical confounders that might affect or are known to affect cognitive task performance, such as neuropsychiatric symptoms (i.e., depression) as well as those clinical measures that were significantly different at baseline between PD patient groups on relevant scales (e.g., PD-related scales). We included the scores at each time point (i.e., study year) as opposed to only baseline scores for all relevant clinical covariates to ensure that reported results control for longitudinal change in these covariates that might otherwise account for cognitive performance differences. PD medications expressed in LEDD (mg/day) were not included in the model, as we did not find any difference in the amount of LEDD or in the trajectory of dopamine-replacement medications between PDnP and PDP patients (please refer to Supplementary Material 1). Treatment for psychosis in PD can include administration of antipsychotics such as clozapine, quetiapine and olanzapine,^41,42^ and antipsychotic treatment has shown to affect cognitive functions.^43,44^ 38 patients (Supplementary Material 1) were reported to take antipsychotics. We carried out analyses both including and after excluding those 38 participants. As the results did not materially differ, here we report the results of analyses including those on antipsychotics; results of analyses excluding them are available upon request. All included covariates of interest were mean-centred for analysis.

### Data availability

Data used in the preparation of this article were obtained [on February 1^st^ 2023] from the Parkinson’s Progression Markers Initiative (PPMI) database (www.ppmi-info.org/access-dataspecimens/download-data), RRID:SCR 006431. For up-to-date information on the study, visit www.ppmi-info.org.

## Results

### Baseline sample characteristics

There were no significant differences across groups on age and sex, however there was a greater prevalence of individuals identifying as “white” compared with other ethnic groups (*P =* 0.008), and PDP patients reported significantly fewer years of educations compared with HC (*P =* 0.024). There were no differences in PD duration, age of diagnosis and age of PD onset between PD groups (all *P* > 0.05) (Table 1). PDP patients reported more rigidity (*P =* 0.032), more severe symptoms on MDS-UPDRS part I (*P =*0.003) and II (*P* < 0.001), more sleepiness (*P =* 0.016), more RBD (*p* < 0.001), more anxiety (*P =* 0.032), and more autonomic symptoms (*P* < 0.001) than PDnP at baseline. Both PD groups did not differ on any cognitive measures at baseline. As expected, PD patients had more severe motor symptoms on all PD-related assessments (MDS-UPDRS, part I, part II and part III, all *P* < 0.05), and greater tremor and rigidity (both *P* < 0.001) compared with those reported by HC. They reported more depressive symptoms compared with HC (*P* < 0.001) and reported more RBD, more anxiety, and more autonomic symptoms (all *P* < 0.001) than HC. PD patients performed significantly worse than HC in the majority of cognitive batteries (all *P* < 0.05) (see Table 1).

**Table 1.**
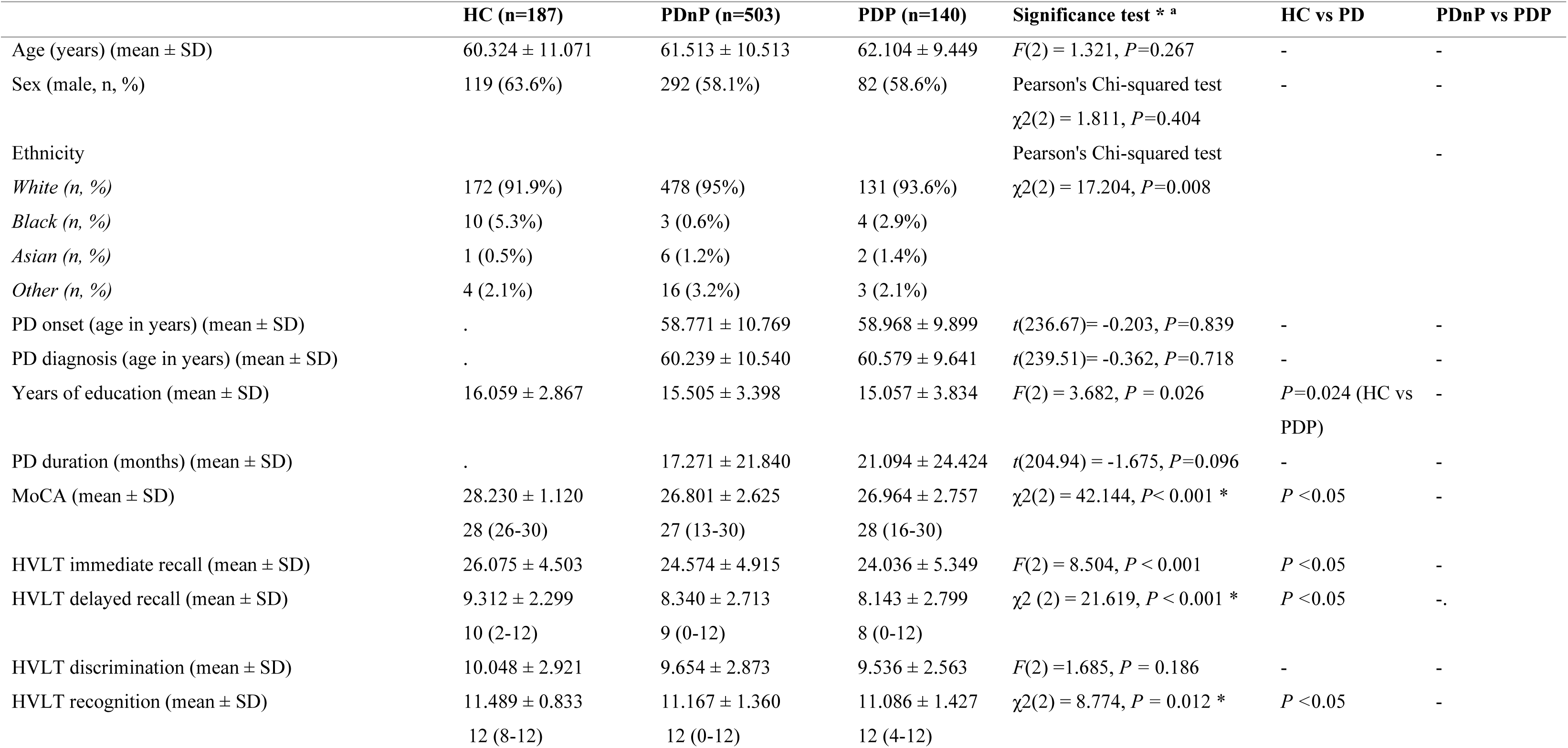
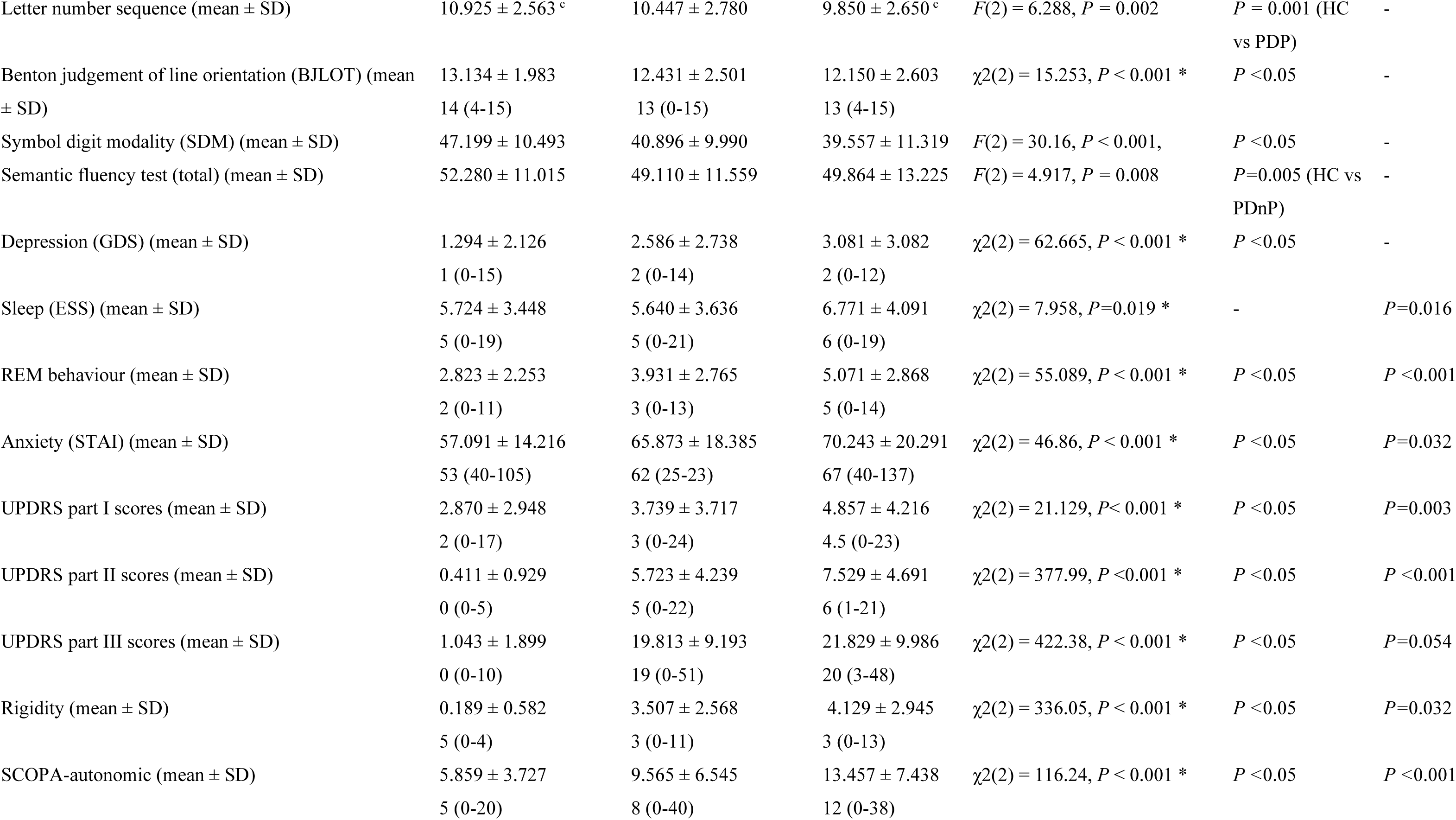
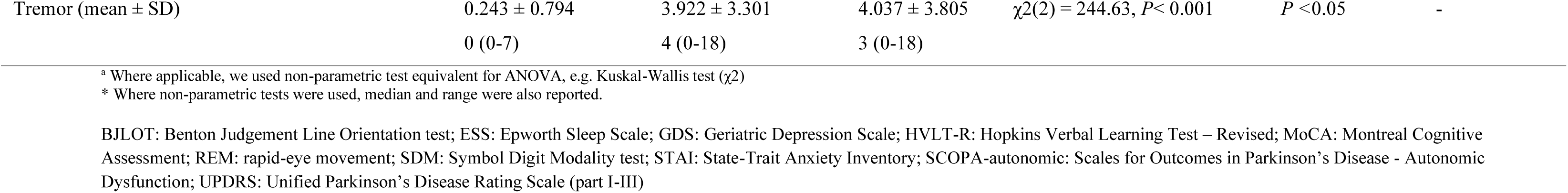
Baseline sample characteristics. Sample characteristics at baseline of the three groups, i.e., PD patients without psychosis (PDnP), PD patients with psychosis (PDP) and healthy controls (HC). PDP psychosis group does not include PD patients who reported psychosis symptoms at baseline. Mean and SDs are reported, unless otherwise specified.

|  | HC (n=187) | PDnP (n=503) | PDP (n=140) | Significance test * <sup>a</sup> | HC vs PD | PDnP vs PDP |
| --- | --- | --- | --- | --- | --- | --- |
| Age (years) (mean ± SD) | 60.324 ± 11.071 | 61.513 ± 10.513 | 62.104 ± 9.449 | $F(2) = 1.321, P=0.267$ | - | - |
| Sex (male, n, %) | 119 (63.6%) | 292 (58.1%) | 82 (58.6%) | Pearson's Chi-squared test<br>$\chi^2(2) = 1.811, P=0.404$ | - | - |
| Ethnicity |  |  |  | Pearson's Chi-squared test |  | - |
| White (n, %) | 172 (91.9%) | 478 (95%) | 131 (93.6%) | $\chi^2(2) = 17.204, P=0.008$ | | |
| Black (n, %) | 10 (5.3%) | 3 (0.6%) | 4 (2.9%) |  |  |  |
| Asian (n, %) | 1 (0.5%) | 6 (1.2%) | 2 (1.4%) |  |  |  |
| Other (n, %) | 4 (2.1%) | 16 (3.2%) | 3 (2.1%) |  |  |  |
| PD onset (age in years) (mean ± SD) | . | 58.771 ± 10.769 | 58.968 ± 9.899 | $t(236.67) = -0.203, P=0.839$ | - | - |
| PD diagnosis (age in years) (mean ± SD) | . | 60.239 ± 10.540 | 60.579 ± 9.641 | $t(239.51) = -0.362, P=0.718$ | - | - |
| Years of education (mean ± SD) | 16.059 ± 2.867 | 15.505 ± 3.398 | 15.057 ± 3.834 | $F(2) = 3.682, P = 0.026$ | $P=0.024$ (HC vs PDP) | - |
| PD duration (months) (mean ± SD) | . | 17.271 ± 21.840 | 21.094 ± 24.424 | $t(204.94) = -1.675, P=0.096$ | - | - |
| MoCA (mean ± SD) | 28.230 ± 1.120 | 26.801 ± 2.625 | 26.964 ± 2.757 | $\chi^2(2) = 42.144, P < 0.001$ * | $P < 0.05$ | - |
|  | 28 (26-30) | 27 (13-30) | 28 (16-30) |  |  |  |
| HVLT immediate recall (mean ± SD) | 26.075 ± 4.503 | 24.574 ± 4.915 | 24.036 ± 5.349 | $F(2) = 8.504, P < 0.001$ | $P < 0.05$ | - |
| HVLT delayed recall (mean ± SD) | 9.312 ± 2.299 | 8.340 ± 2.713 | 8.143 ± 2.799 | $\chi^2(2) = 21.619, P < 0.001$ * | $P < 0.05$ | - |
|  | 10 (2-12) | 9 (0-12) | 8 (0-12) |  |  |  |
| HVLT discrimination (mean ± SD) | 10.048 ± 2.921 | 9.654 ± 2.873 | 9.536 ± 2.563 | $F(2) = 1.685, P = 0.186$ | - | - |
| HVLT recognition (mean ± SD) | 11.489 ± 0.833 | 11.167 ± 1.360 | 11.086 ± 1.427 | $\chi^2(2) = 8.774, P = 0.012$ * | $P < 0.05$ | - |
|  | 12 (8-12) | 12 (0-12) | 12 (4-12) |  |  |  |
| Letter number sequence (mean ± SD) | 10.925 ± 2.563<sup>c</sup> | 10.447 ± 2.780 | 9.850 ± 2.650<sup>c</sup> | $F(2) = 6.288, P = 0.002$ | $P = 0.001$ (HC vs PDP) | - |
| Benton judgement of line orientation (BJLOT) (mean ± SD) | 13.134 ± 1.983<br>14 (4-15) | 12.431 ± 2.501<br>13 (0-15) | 12.150 ± 2.603<br>13 (4-15) | $\chi^2(2) = 15.253, P < 0.001 *$ | $P < 0.05$ | - |
| Symbol digit modality (SDM) (mean ± SD) | 47.199 ± 10.493 | 40.896 ± 9.990 | 39.557 ± 11.319 | $F(2) = 30.16, P < 0.001,$ | $P < 0.05$ | - |
| Semantic fluency test (total) (mean ± SD) | 52.280 ± 11.015 | 49.110 ± 11.559 | 49.864 ± 13.225 | $F(2) = 4.917, P = 0.008$ | $P = 0.005$ (HC vs PDnP) | - |
| Depression (GDS) (mean ± SD) | 1.294 ± 2.126<br>1 (0-15) | 2.586 ± 2.738<br>2 (0-14) | 3.081 ± 3.082<br>2 (0-12) | $\chi^2(2) = 62.665, P < 0.001 *$ | $P < 0.05$ | - |
| Sleep (ESS) (mean ± SD) | 5.724 ± 3.448<br>5 (0-19) | 5.640 ± 3.636<br>5 (0-21) | 6.771 ± 4.091<br>6 (0-19) | $\chi^2(2) = 7.958, P = 0.019 *$ | - | $P = 0.016$ |
| REM behaviour (mean ± SD) | 2.823 ± 2.253<br>2 (0-11) | 3.931 ± 2.765<br>3 (0-13) | 5.071 ± 2.868<br>5 (0-14) | $\chi^2(2) = 55.089, P < 0.001 *$ | $P < 0.05$ | $P < 0.001$ |
| Anxiety (STAI) (mean ± SD) | 57.091 ± 14.216<br>53 (40-105) | 65.873 ± 18.385<br>62 (25-23) | 70.243 ± 20.291<br>67 (40-137) | $\chi^2(2) = 46.86, P < 0.001 *$ | $P < 0.05$ | $P = 0.032$ |
| UPDRS part I scores (mean ± SD) | 2.870 ± 2.948<br>2 (0-17) | 3.739 ± 3.717<br>3 (0-24) | 4.857 ± 4.216<br>4.5 (0-23) | $\chi^2(2) = 21.129, P < 0.001 *$ | $P < 0.05$ | $P = 0.003$ |
| UPDRS part II scores (mean ± SD) | 0.411 ± 0.929<br>0 (0-5) | 5.723 ± 4.239<br>5 (0-22) | 7.529 ± 4.691<br>6 (1-21) | $\chi^2(2) = 377.99, P < 0.001 *$ | $P < 0.05$ | $P < 0.001$ |
| UPDRS part III scores (mean ± SD) | 1.043 ± 1.899<br>0 (0-10) | 19.813 ± 9.193<br>19 (0-51) | 21.829 ± 9.986<br>20 (3-48) | $\chi^2(2) = 422.38, P < 0.001 *$ | $P < 0.05$ | $P = 0.054$ |
| Rigidity (mean ± SD) | 0.189 ± 0.582<br>5 (0-4) | 3.507 ± 2.568<br>3 (0-11) | 4.129 ± 2.945<br>3 (0-13) | $\chi^2(2) = 336.05, P < 0.001 *$ | $P < 0.05$ | $P = 0.032$ |
| SCOPA-autonomic (mean ± SD) | 5.859 ± 3.727<br>5 (0-20) | 9.565 ± 6.545<br>8 (0-40) | 13.457 ± 7.438<br>12 (0-38) | $\chi^2(2) = 116.24, P < 0.001 *$ | $P < 0.05$ | $P < 0.001$ |
| Tremor (mean ± SD) | 0.243 ± 0.794 | 3.922 ± 3.301 | 4.037 ± 3.805 | $\chi^2(2) = 244.63, P < 0.001$ | $P < 0.05$ | - |
|  | 0 (0-7) | 4 (0-18) | 3 (0-18) |  |  |  |

<sup>a</sup> Where applicable, we used non-parametric test equivalent for ANOVA, e.g. Kuskal-Wallis test ( $\chi^2$ )
\* Where non-parametric tests were used, median and range were also reported.
BJLOT: Benton Judgement Line Orientation test; ESS: Epworth Sleep Scale; GDS: Geriatric Depression Scale; HVLT-R: Hopkins Verbal Learning Test – Revised; MoCA: Montreal Cognitive Assessment; REM: rapid-eye movement; SDM: Symbol Digit Modality test; STAI: State-Trait Anxiety Inventory; SCOPA-autonomic: Scales for Outcomes in Parkinson's Disease - Autonomic Dysfunction; UPDRS: Unified Parkinson's Disease Rating Scale (part I-III)

**Table 2.** PD psychosis patients. PD patients who developed psychosis from year 1 to year 5 of the PPMI study, followed by the number (n) and percentage (%) of how many reported score 1-3 of the MDS-UPDRS part 1 item 1.2. A score of 0 = normal, 1 = minor illusions and non-formed hallucinations, 2 = formed hallucinations, 3 = formed hallucinations without insight, 4 = psychosis with delusions and hallucinations.

| Study year (PD patients with psychosis, n) | Score = 1 | Score = 2 | Score = 3 |
| --- | --- | --- | --- |
| Year 1 (n=37) | 34 (91.9%) | 3 (8.1%) | 0 (0) |
| Year 2 (n=43) | 35 (81.4%) | 7 (16.3%) | 1 (2.3%) |
| Year 3 (n=48) | 38 (79.2%) | 9 (18.8%) | 1 (2.1%) |
| Year 4 (n=49) | 44 (89.8%) | 3 (6.1%) | 2 (4.1%) |
| Year 5 (n=51) | 41 (80.4%) | 8 (15.7%) | 2 (3.9%) |

### Trajectories of cognitive performance across groups

#### HC vs PDP and PDnP

Table 3 and Table 4 report the results from the unadjusted and adjusted analyses respectively. Figure 2 reports the predicted values of task performances for all cognitive domains. As expected, there was a significant main effect of group across all time points (i.e., year 0 to year 5) showing a worse performance of PD groups over HC across HVLT-R tests (i.e., immediate recall, and delayed recall, both *P <*0.001, and recognition, *P=*0.006 (PDnP), *P=*0.007 (PDP)). Results showed a worse performance of PD patients compared with HC on semantic fluency (PDnP vs HC, *P=*0.006), LNS (PDP vs HC, *P=*0.015), SDM, and MoCA (all *P <*0.001). PD patients also reported a significantly worse trajectory of cognitive decline compared with HC across all tests, except for recognition (PDnP, *P=*0.208; PDP, *P=*0.221), and delayed recall (PDP, *P=*0.08) (see Table 3). After controlling for covariates of interest, the longitudinal trajectories of task performance across the majority of cognitive measures remained significantly worse in PD patients compared to HC (Supplementary Material 3, eTable5).

**Figure 2.**
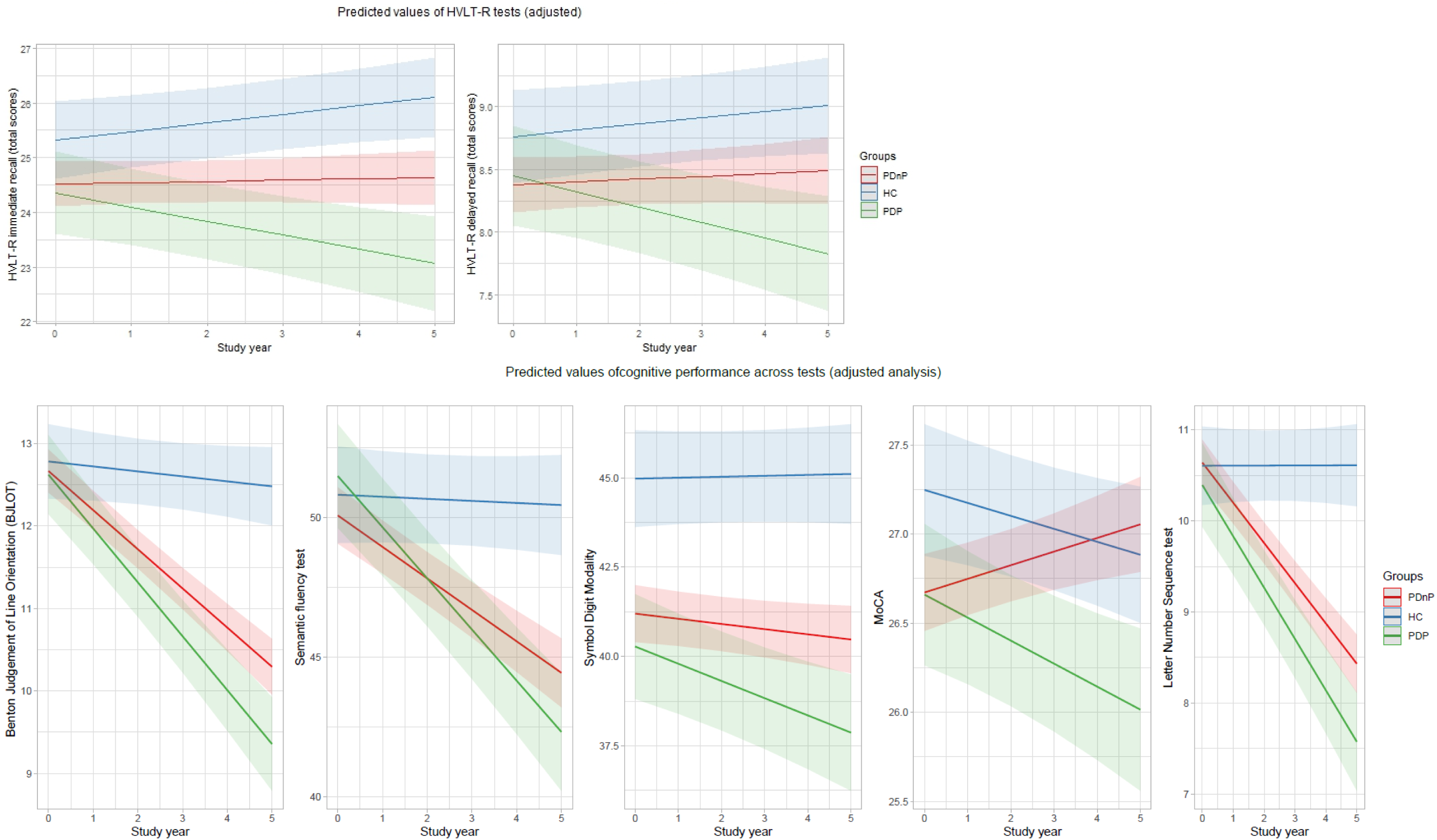
Predictive values from the adjusted analyses. Predicted values (with associated 95% confidence intervals) from the analysis controlled for depression, sleepiness, REM behaviour sleep disorder and motor symptom severity. Study years (from baseline, year 0 to year 5) on the x axis, and total scores of cognitive tests on the y axis.

**Table 3.**
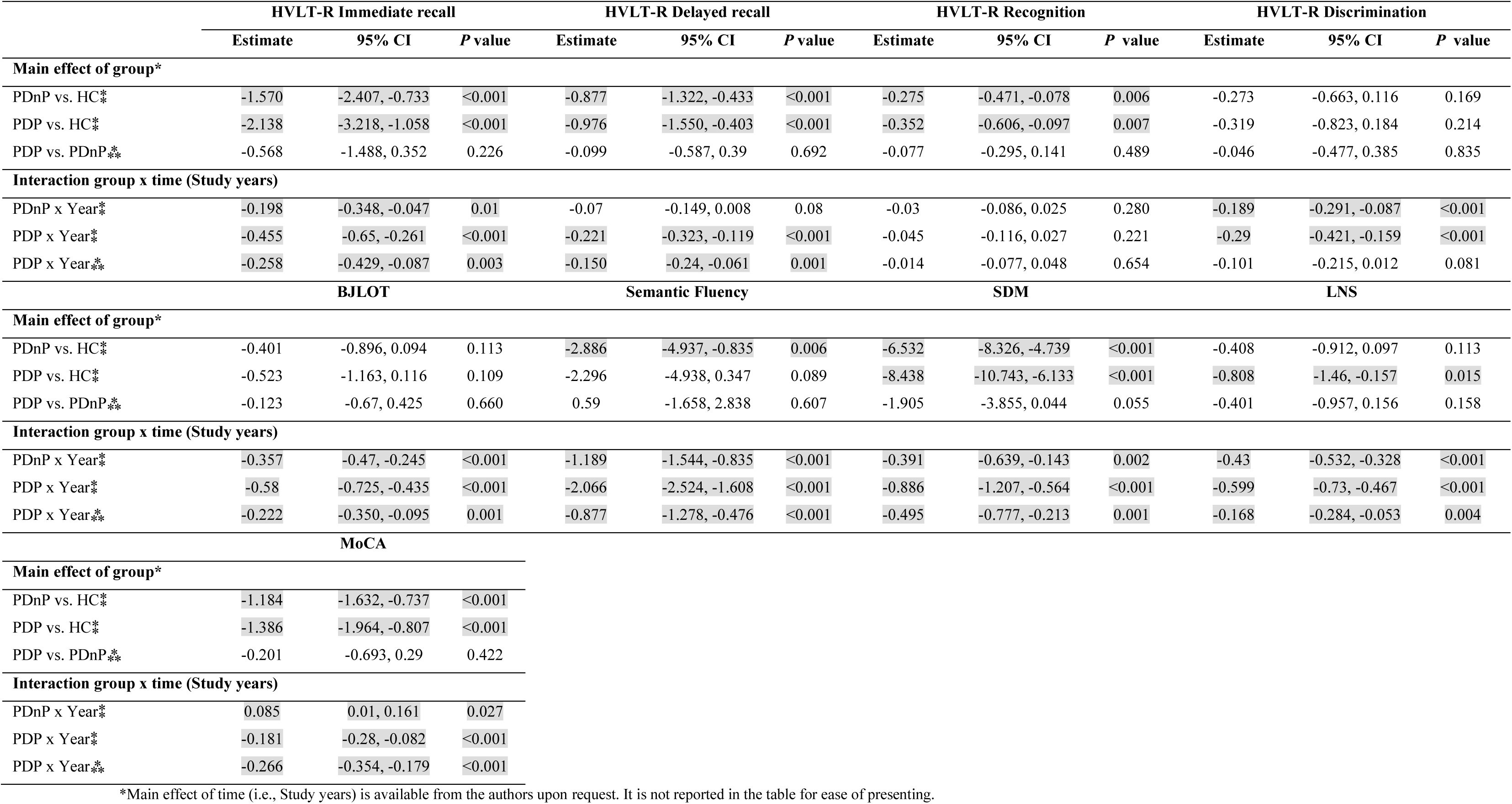
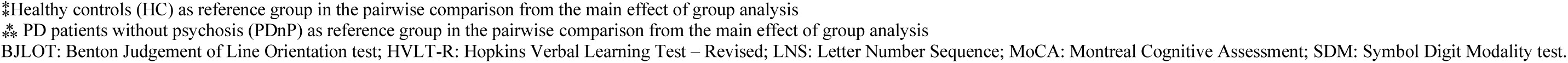
Results from unadjusted analysis. Unadjusted group comparison of cognitive performance trajectory between HC, PDnP, and PDP.

**Table 4.**
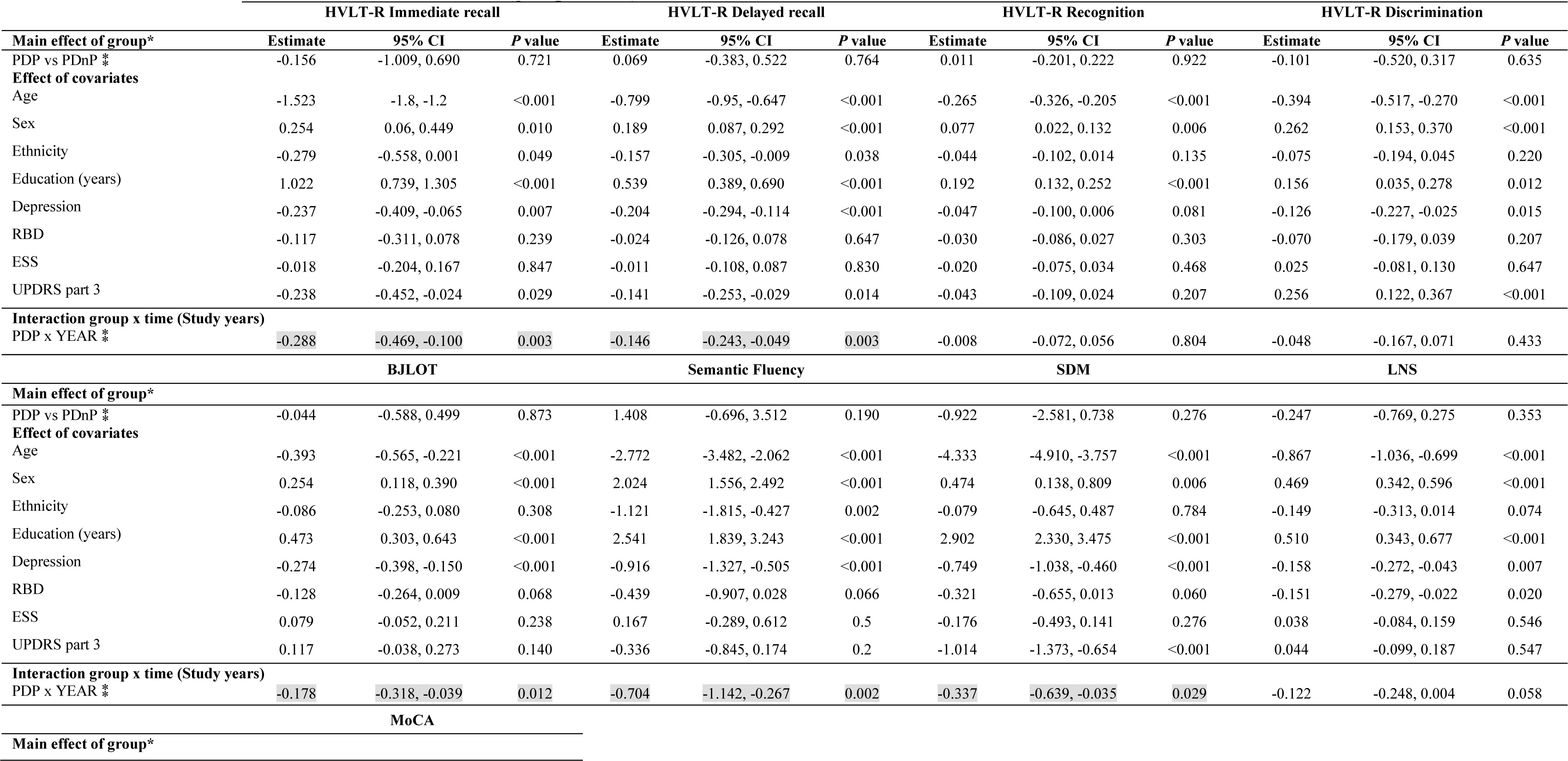
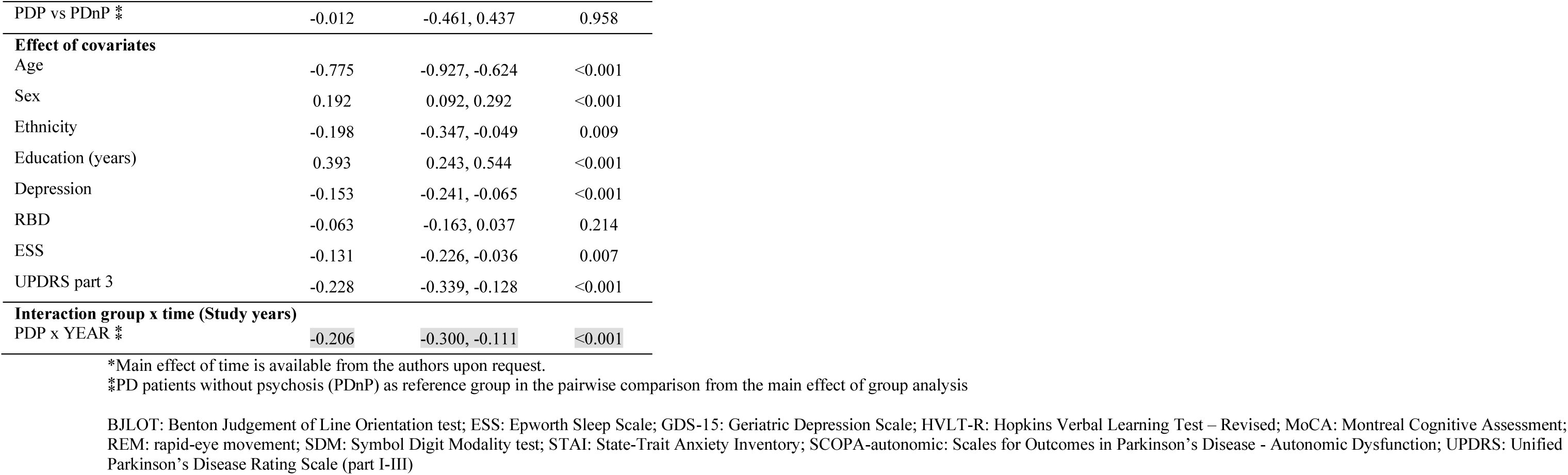
Results from the adjusted analysis. Adjusted comparison of cognitive performance trajectories: For ease of presentation, only PDP vs PDnP comparisons reported here (for comparison between HC and PD group, please refer to the Supplementary Material 3, eTable5 A and B). Analyses were adjusted for depression, sleepiness, REM behaviour sleep disorder, and motor symptoms. Highlighted in grey, significant results pertaining to the main effect and interaction (group * time).

|  | HVLt-R Immediate recall |  |  | HVLt-R Delayed recall |  |  | HVLt-R Recognition |  |  | HVLt-R Discrimination |  |  |
| --- | --- | --- | --- | --- | --- | --- | --- | --- | --- | --- | --- | --- |
| Main effect of group* | Estimate | 95% CI | P value | Estimate | 95% CI | P value | Estimate | 95% CI | P value | Estimate | 95% CI | P value |
| PDP vs PDnP ‡ | -0.156 | -1.009, 0.690 | 0.721 | 0.069 | -0.383, 0.522 | 0.764 | 0.011 | -0.201, 0.222 | 0.922 | -0.101 | -0.520, 0.317 | 0.635 |
| Effect of covariates |  |  |  |  |  |  |  |  |  |  |  |  |
| Age | -1.523 | -1.8, -1.2 | <0.001 | -0.799 | -0.95, -0.647 | <0.001 | -0.265 | -0.326, -0.205 | <0.001 | -0.394 | -0.517, -0.270 | <0.001 |
| Sex | 0.254 | 0.06, 0.449 | 0.010 | 0.189 | 0.087, 0.292 | <0.001 | 0.077 | 0.022, 0.132 | 0.006 | 0.262 | 0.153, 0.370 | <0.001 |
| Ethnicity | -0.279 | -0.558, 0.001 | 0.049 | -0.157 | -0.305, -0.009 | 0.038 | -0.044 | -0.102, 0.014 | 0.135 | -0.075 | -0.194, 0.045 | 0.220 |
| Education (years) | 1.022 | 0.739, 1.305 | <0.001 | 0.539 | 0.389, 0.690 | <0.001 | 0.192 | 0.132, 0.252 | <0.001 | 0.156 | 0.035, 0.278 | 0.012 |
| Depression | -0.237 | -0.409, -0.065 | 0.007 | -0.204 | -0.294, -0.114 | <0.001 | -0.047 | -0.100, 0.006 | 0.081 | -0.126 | -0.227, -0.025 | 0.015 |
| RBD | -0.117 | -0.311, 0.078 | 0.239 | -0.024 | -0.126, 0.078 | 0.647 | -0.030 | -0.086, 0.027 | 0.303 | -0.070 | -0.179, 0.039 | 0.207 |
| ESS | -0.018 | -0.204, 0.167 | 0.847 | -0.011 | -0.108, 0.087 | 0.830 | -0.020 | -0.075, 0.034 | 0.468 | 0.025 | -0.081, 0.130 | 0.647 |
| UPDRS part 3 | -0.238 | -0.452, -0.024 | 0.029 | -0.141 | -0.253, -0.029 | 0.014 | -0.043 | -0.109, 0.024 | 0.207 | 0.256 | 0.122, 0.367 | <0.001 |
| Interaction group x time (Study years) |  |  |  |  |  |  |  |  |  |  |  |  |
| PDP x YEAR ‡ | -0.288 | -0.469, -0.100 | 0.003 | -0.146 | -0.243, -0.049 | 0.003 | -0.008 | -0.072, 0.056 | 0.804 | -0.048 | -0.167, 0.071 | 0.433 |
|  | BJLOT |  |  | Semantic Fluency |  |  | SDM |  |  | LNS |  |  |
| Main effect of group* |  |  |  |  |  |  |  |  |  |  |  |  |
| PDP vs PDnP ‡ | -0.044 | -0.588, 0.499 | 0.873 | 1.408 | -0.696, 3.512 | 0.190 | -0.922 | -2.581, 0.738 | 0.276 | -0.247 | -0.769, 0.275 | 0.353 |
| Effect of covariates |  |  |  |  |  |  |  |  |  |  |  |  |
| Age | -0.393 | -0.565, -0.221 | <0.001 | -2.772 | -3.482, -2.062 | <0.001 | -4.333 | -4.910, -3.757 | <0.001 | -0.867 | -1.036, -0.699 | <0.001 |
| Sex | 0.254 | 0.118, 0.390 | <0.001 | 2.024 | 1.556, 2.492 | <0.001 | 0.474 | 0.138, 0.809 | 0.006 | 0.469 | 0.342, 0.596 | <0.001 |
| Ethnicity | -0.086 | -0.253, 0.080 | 0.308 | -1.121 | -1.815, -0.427 | 0.002 | -0.079 | -0.645, 0.487 | 0.784 | -0.149 | -0.313, 0.014 | 0.074 |
| Education (years) | 0.473 | 0.303, 0.643 | <0.001 | 2.541 | 1.839, 3.243 | <0.001 | 2.902 | 2.330, 3.475 | <0.001 | 0.510 | 0.343, 0.677 | <0.001 |
| Depression | -0.274 | -0.398, -0.150 | <0.001 | -0.916 | -1.327, -0.505 | <0.001 | -0.749 | -1.038, -0.460 | <0.001 | -0.158 | -0.272, -0.043 | 0.007 |
| RBD | -0.128 | -0.264, 0.009 | 0.068 | -0.439 | -0.907, 0.028 | 0.066 | -0.321 | -0.655, 0.013 | 0.060 | -0.151 | -0.279, -0.022 | 0.020 |
| ESS | 0.079 | -0.052, 0.211 | 0.238 | 0.167 | -0.289, 0.612 | 0.5 | -0.176 | -0.493, 0.141 | 0.276 | 0.038 | -0.084, 0.159 | 0.546 |
| UPDRS part 3 | 0.117 | -0.038, 0.273 | 0.140 | -0.336 | -0.845, 0.174 | 0.2 | -1.014 | -1.373, -0.654 | <0.001 | 0.044 | -0.099, 0.187 | 0.547 |
| Interaction group x time (Study years) |  |  |  |  |  |  |  |  |  |  |  |  |
| PDP x YEAR ‡ | -0.178 | -0.318, -0.039 | 0.012 | -0.704 | -1.142, -0.267 | 0.002 | -0.337 | -0.639, -0.035 | 0.029 | -0.122 | -0.248, 0.004 | 0.058 |
|  | MoCA |  |  |  |  |  |  |  |  |  |  |  |
| Main effect of group* |  |  |  |  |  |  |  |  |  |  |  |  |

|  |  |  |  |
| --- | --- | --- | --- |
| PDP vs PDnP * | -0.012 | -0.461, 0.437 | 0.958 |
| **Effect of covariates** |  |  |  |
| Age | -0.775 | -0.927, -0.624 | <0.001 |
| Sex | 0.192 | 0.092, 0.292 | <0.001 |
| Ethnicity | -0.198 | -0.347, -0.049 | 0.009 |
| Education (years) | 0.393 | 0.243, 0.544 | <0.001 |
| Depression | -0.153 | -0.241, -0.065 | <0.001 |
| RBD | -0.063 | -0.163, 0.037 | 0.214 |
| ESS | -0.131 | -0.226, -0.036 | 0.007 |
| UPDRS part 3 | -0.228 | -0.339, -0.128 | <0.001 |
| **Interaction group x time (Study years)** |  |  |  |
| PDP x YEAR * | -0.206 | -0.300, -0.111 | <0.001 |

\*Main effect of time is available from the authors upon request.
\*PD patients without psychosis (PDnP) as reference group in the pairwise comparison from the main effect of group analysis
BJLOT: Benton Judgement of Line Orientation test; ESS: Epworth Sleep Scale; GDS-15: Geriatric Depression Scale; HVLT-R: Hopkins Verbal Learning Test – Revised; MoCA: Montreal Cognitive Assessment; REM: rapid-eye movement; SDM: Symbol Digit Modality test; STAI: State-Trait Anxiety Inventory; SCOPA-autonomic: Scales for Outcomes in Parkinson's Disease - Autonomic Dysfunction; UPDRS: Unified Parkinson's Disease Rating Scale (part I-III)

#### PDP and PDnP

In terms of our main hypothesis of interest, unadjusted analyses showed a significant difference in the trajectory of cognitive performance between PDP and PDnP across most measures (i.e., interaction of group * time) over the 5-year follow-up period. PDP patients showed a significantly worse trajectory over time compared to PDnP patients across all cognitive tasks, except recognition (*P=*0.610), and discrimination (*P=*0.081). After adjusting for age, sex, ethnicity, years of education, depression, sleepiness, RBD, and severity of motor symptoms, the longitudinal trajectories remained significantly different between PDP and PDnP for most of the cognitive measures, namely HVLT-R immediate (b=-0.288, 95% CI -0.452, -0.100, *P=*0.003) and delayed recall (b=-0.146, 95% CI -0.243, -0.049, *P=*0.003), BJLOT (b=-0.178, 95% CI -0.318, -0.039, *P=*0.012), semantic fluency (b=-0.704, 95% CI, -1.142, -0.267, *P=*0.002), SDM (b=-0.337, 95% CI -0.639, -0.035, *P=*0.029), and MoCA (b=-0.206, 95% CI, -0.300, -0.111, *P <*0.001). There was also a trend towards a difference in longitudinal trajectory of LNS between PDP and PDnP patients (*P=*0.058).

## Discussion

In this study, we investigated the trajectory of cognitive function in PD patients with and without psychosis and healthy control individuals. As expected, and in line with previous reports,^13,14,21^ both groups of PD patients showed a progressive worsening of performance across a range of cognitive domains, specifically, semantic aspects of language, processing speed, memory, visuo-spatial and general cognitive abilities, during the early years of their clinical course compared to age and sex-matched healthy individuals. These differences were evident even after controlling for sociodemographic factors (i.e., age, sex, ethnicity, years of education) as well as the longitudinal course of clinical factors that may potentially account for these different trajectories. Consistent with our main overarching hypothesis of interest, we also found that PD psychosis participants showed a progressively greater deterioration over time in memory (i.e., HVLT-R immediate, and delayed recall), processing speed (Symbol Digit Modality), language (semantic fluency), visuo-spatial abilities (Benton Judgement of Line Orientation) and general global cognition (MoCA) compared with PD participants without psychosis. These effects were evident even after controlling for the potential confounding effects of age, sex, ethnicity and years of education as well as the longitudinal course of depression, sleepiness, RBD and severity of motor symptoms over the same period in those participants. Such a differential trajectory of worsening was not evident for cognitive domains such as recognition and discrimination components of memory, and working memory, namely manipulation (as measured with the Letter Number Sequence). Our findings are broadly in line with previous meta-analytic evidence ^10,11^ from cross-sectional studies indicating impairments in global cognition, visuo-spatial, language, processing speed as well as in sub-domains such as manipulation, immediate recall, category-based fluency in PD patients with psychosis compared to those without. However, as these analyses were based on cross-sectional studies, the longitudinal course of these impairments remained unclear. To the best of our knowledge, only a small number of studies to date have investigated the longitudinal course of cognition in PD psychosis. While Morgante et al. ^13^ reported lower global cognition as indexed by MMSE scores at 2-year follow up in PD patients who developed psychosis, Goetz et al. ^15^ did not find a similar effect. However, Muller et al. ^14^ found poorer performance in processing speed and executive function at follow up in PD patients who developed visual hallucinations (n=18) compared to those who did not (n=15). In a large prospective cohort of PD participants (n=676), here we extend these findings to show that compared to healthy volunteers (n=187), there is a progressive impairment in a number of domains of cognition in people with PD without psychosis at baseline and that this is even greater in people who go on to develop psychosis over the first 5 years following presentation to clinical services, despite both groups of PD participants being comparable in terms of cognitive task performance at the initiation of cohort. Further, this worsening trajectory of cognition could not be explained by group differences in socio-demographic factors or longitudinal course of potential clinical confounders. For the two most affected domains of semantic fluency and processing speed (Symbol Digit Modality) that we observed in this study, this translates to patients with PD psychosis scoring an additional 0.7 words and 0.337 points worse respectively than PD patients without psychosis for each additional year of illness from baseline. The association between psychosis in people with PD and development of dementia is well-recognised.^45^ Although, we excluded from analysis participants who developed dementia during the 5-year follow-up period of the study, we cannot be certain that some of the group level cognitive impairments observed in PDP participants were not early indicators of the impending transition to dementia. On average, task performance in the PDP group was at least 1 standard deviation below that in the HC group in more than one cognitive domain, particularly during the latter period of follow-up of the PPMI cohort. Hence, it is possible that many of them may have met the MDS PD-mild cognitive impairment (PD-MCI) diagnostic criteria.^45,46^ Results presented here, therefore underscore the need to investigate the relationship between progressive decline in these cognitive domains in people with PDP and eventual clinical diagnosis of dementia in future studies.

Interestingly, our findings highlight the greater progressive worsening in semantic fluency, a measure of semantic aspects of language, in PD patients who develop psychosis compared to those who do not, which is consistent with previous evidence of language deficits in hallucinating PD patients.^5,6^ Semantic decline as revealed by similar tests to those used in the PPMI cohort is a core feature of fronto-temporal dementia and other neurodegenerative conditions.^47–49^ Consistent with our results, impaired baseline semantic fluency ^50,51^ in conjunction with suboptimal performance in symbol digit modality, and recall (as measured with the HVLT-R) tasks^52^ have been shown to predict cognitive decline in PD patients. Semantic deficits in these conditions are associated with AD neuropathology, primarily in the anterior temporal lobe. Although the underlying neuropathology differs, the same anterior temporal cortex region, particularly the amygdala, is associated with VH in the context of PD or Dementia with Lewy Bodies.^53,54^ Our findings show that PD patients who develop psychosis report a greater decline in semantic fluency compared to PD patients without psychosis. The initial changes observed in PD psychosis involve Lewy Body neuropathology within the anterior temporal lobe ^53^ suggesting that a combination of Lewy Body and AD neuropathology may be responsible for both semantic fluency decline and onset of PD psychosis. We also observed greater progressive worsening in PDP compared to PDnP in processing speed, which is consistent with the suggestion that visual hallucinations in PD may be a result of dysfunctional attentional processing due to impaired attentional networks such as the ventral and dorsal attentional networks.^55–57^ As we have observed declining performance across visuo-spatial abilities, overall general cognitive abilities (as measured with the MoCA) and memory (namely immediate and delayed recall), attributing the emergence of psychosis in PD solely to dysfunctions and altered connections between dorsal and ventral attention networks might oversimply the complex brain mechanisms involved. Notwithstanding the relevance of the current model of visual hallucinations proposed by Collerton et al.^57^, the present work may seem to suggest that visual hallucination, or more generally psychosis, may not solely be due to visual processing deficits and impairments in attention. Semantic fluency or semantic loss should be considered as part of this model, as a symptom in PD psychosis and included in the model for a more comprehensive understanding. In line with this, it is important to remember that we do not know yet if semantic deficits preceded visual deficits or vice versa, therefore this needs to be tested. Similarly, Collerton et al. ^57^ and Shine et al. ^56,58^ models emphasise the involvement of dorsal and ventral networks, as well as the Default Mode Network in PD psychosis. Our results suggest including the anterior temporal lobe and semantic aspects of language in the model. Collectively, this warrants more examination to obtain insights in temporal regions potentially affected in PD psychosis.

Given the prevalence of psychosis in PD is 20-30%,^59^ our findings further highlight the urgency of prioritising treatment for cognitive impairments. Hallucinations and cognitive deficits may lead to poor quality of life,^60^ namely poor verbal fluency is associated with deteriorating health and increased care burden in PD.^61^ Preventative measures such as cholinesterase inhibitors,^62,63^ especially Rivastigmine, are commonly used in PD for psychosis and cognitive impairment ^64^ and have demonstrated benefits in the domains of praxis, memory and executive functioning in neurodegenerative disorders.^65,66^ Notably, our findings align with Aarsland et al.’s ^45^ report, indicating that the observed longitudinal changes in performance across domains in PD psychosis might be leading to more severe cognitive impairments and then dementia. Our results suggest a parallel observation proposing the inclusion of language-based tests (i.e., semantic category-based fluency), along with assessments like the intersecting pentagon (see for example ^51^), for enhanced screening to identify dementia progression in PD, especially in PD psychosis. Lastly, we also observed a significantly worse trajectory of performance in PDnP patients compared with HC across two cognitive tasks, after adjusting for covariates of interest. Although we had predicted that this would be evident across all cognitive domains, we found that this difference was significant only for Symbol Digit Modality (processing speed) and MoCA. This is an expected result as healthy individuals generally report less severe cognitive decline compared to individuals with neurodegenerative diseases.^67^

The present study benefits from the largest longitudinal sample investigating this question to date and data on a comprehensive set of cognitive tasks/domains as well as potential confounders. Further, we employed a mixed-effect analytic approach that allowed us to investigate whether the longitudinal course of performance changes differ between the participant groups, while accounting for the correlated nature of the repeated measurements in each individual. However, there are also certain limitations to note. One of the limitations of the present endeavour may relate to how we have operationalised the definition of PD psychosis. We used a classification method derived from a previous study ^18^ but grouped people with milder symptoms such as visual illusions with those experiencing symptoms at the more severe end of the spectrum. Arguably, this approach of combining those with milder and more severe symptoms may have reduced the sensitivity of our analyses to detect any group differences. Nevertheless, we were able to detect significant differences between PDP and PDnP in the longitudinal course of cognitive task performance. We used the MDS-UPDRS part 1 “hallucination/psychosis” question to classify PD patients into PD with and without psychosis. However, this question, to some extent, lacks specificity in classifying patients into distinct psychosis symptom categories. For instance, a score of 4 indicates presence of paranoid thoughts yet it does not distinctly delineate whether patients exhibits both hallucinations and delusions, delusions alone or delusions resulting from presence of hallucinations. No other measure employed in the PPMI study can provide such information, therefore we were not able to clearly define groups based different symptoms of psychosis, and future research should consider applying different assessments such as Scale for Assessment of Positive Symptoms for PD (SAPS-PD) ^68^ or the Neuropsychiatric Inventory (NPI) ^69^ to examine cognitive trajectory in groups of PDP patients with different symptoms. It is also worth noting that we did not investigate the cognitive trajectories from baseline until immediately before the onset of psychotic symptoms. Instead, we analysed task performance scores from all those who had developed psychosis within the 5-year follow-up period even after the onset of their psychotic symptoms, as long as they did not have such symptoms at cohort initiation (or time 0). Therefore, some of the greater worsening of longitudinal course of cognition in PDP compared to PDnP that we report here may reflect the fact that some of the PDP patients were already psychotic when they performed the cognitive tasks. We employed this approach in order to ensure that we could make maximal use of the data. Including only cognitive task performance scores until before psychosis onset would have resulted in a substantially reduced sample size in the PDP group. Future studies are therefore warranted to investigate whether a similar pattern of worse longitudinal decline in PDP compared to PDnP is observed even when considering cognitive task performance until before the onset of psychotic symptoms. Another potential limitation relates to the inclusion of those on antipsychotic medications, well-known to affect cognitive performance,^43,44^ in our analysis sample. However, sensitivity analyses (available from the authors upon request) excluding those on antipsychotic medications did not change the direction or pattern of results and hence we have reported results from the complete sample. Evidence from AD studies indicates that atypical antipsychotics could contributed to greater cognitive decline (as measured with the Mini-Mental State Examination, MMSE) compared to placebo in a sample of 421 patients with AD.^70^ Conversely, our results seem to suggest that the use of antipsychotics does not modulate the longitudinal changes across the 5 years of the PPMI study in cognitive performance in PD patients with psychosis. Lastly, it is worth noting the involvement of genetic influences in cognitive decline in PD, specifically glucocerebrosidase (GBA).^71,72^ GBA-related PD is associated with greater cognitive decline in visuo-spatial and executive functions and increased risk of non-motor symptoms such as depression, sleep-related disorders and autonomic dysfunctions.^73^ Future studies should consider exploring the association between genetic influences in PD and the presence of psychosis symptoms.

In conclusion, we examined the trajectory of cognitive performance in PD patients with psychosis and compared it with that of PD patients without psychosis. PD psychosis patients showed severe deterioration across most domains, especially semantic aspects of language and processing speed, independent of socio-demographics, neuropsychiatric symptoms, and motor symptom severity. These findings indicate a potential role of semantic processing in PD psychosis, which should be further examined. Semantic and other language impairments could pave the way to a deeper understanding of psychosis in PD as well as lead to more targeted treatment.

**CRediT authors’ contributions:**

*Conceptualisation:* Sara Pisani, Sagnik Bhattacharyya

*Methodology:* Sara Pisani, Sagnik Bhattacharyya

*Investigation:* Sara Pisani, Latha Velayudhan, Dominic ffytche, Sagnik Bhattacharyya

*Data curation:* Sara Pisani, Luca Gosse

*Formal analysis:* Sara Pisani, Sagnik Bhattacharyya

*Visualisation*: Sara Pisani, Sagnik Bhattacharyya

*Funding acquisition*: Sagnik Bhattacharyya, Latha Velayudhan, Dominic ffytche, Dag Aarsland, Kallol Ray Chaudhuri, Clive Ballard

*Writing (original draft):* Sara Pisani, Sagnik Bhattacharyya, Latha Velayudhan

*Writing (review & editing):* All authors

*Supervision*: Latha Velayudhan, Dominic ffytche, Sagnik Bhattacharyya

## Acknowledgement

Data used in this work were obtained from the Parkinson’s Progression Markers Initiative database (http://www.ppmi-info.org/). PPMI – a public-private partnership – is funded by the Michael J. Fox Foundation for Parkinson’s Research and funding partners, including 4D Pharma, Abbvie, AcureX, Allergan, Amathus Therapeutics, Aligning Science Across Parkinson’s, AskBio, Avid Radiopharmaceuticals, BIAL, Biogen, Biohaven, BioLegend, BlueRock Therapeutics, Bristol-Myers Squibb, Calico Labs, Celgene, Cerevel Therapeutics, Coave Therapeutics, DaCapo Brainscience, Denali, Edmond J. Safra Foundation, Eli Lilly, Gain Therapeutics, GE HealthCare, Genentech, GSK, Golub Capital, Handl Therapeutics, Insitro, Janssen Neuroscience, Lundbeck, Merck, Meso Scale Discovery, Mission Therapeutics, Neurocrine Biosciences, Pfizer, Piramal, Prevail Therapeutics, Roche, Sanofi, Servier, Sun Pharma Advanced Research Company, Takeda, Teva, UCB, Vanqua Bio, Verily, Voyager Therapeutics, the Weston Family Foundation and Yumanity Therapeutics. For up-to-date information on the study, visit www.ppmi-info.org.

## Ethical approval disclosure

The authors of this work confirm that all sites involved in the PPMI study received ethical approval from the ethical standards committees overseeing human experiments before commencing the study. Written informed consent for research was obtained from all participants in the study.

