## Supplementary Material for "Parkinson’s disease psychosis associated with accelerated multi-domain cognitive decline"

### Supplementary Material 1

*Levodopa equivalent daily dose (LEDD)*

PD medications were recorded across all study visits. In brief, we have downloaded the most recent dataset in February 2023 and have extracted dopamine-replacement medication information for 200 new patients which were not included in the curated data prepared by the PPMI study team. PD medications were converted into Levodopa Equivalent Daily Dose (i.e., LEDD) according to Tomlison et al. [1] recommendations. The PPMI study team has reported in a detailed document the different conversion factors, and the procedures. Please visit their website www.ppmi-info.org/study-design for more information. eTable1 shows of the amount of LEDD for each patient's group from year 1 to year 5 follow up.

There were no overall differences between PDnP and PDP (*P*=0.140) in LEDD, but there was a significant difference between PDnP and PDP at baseline (t=3.807, b=369.777, *P*<0.001). LEDD amount increased significantly across the years, on similar degree for all three patients' groups (t=13.493, b=91.837, *P*<0.001) (eFigure 1).

**eTable1.** Means and SDs of LEDD for PD patients (PDnP, PDP and PDP at baseline).

| Study year | Group (n, number of patients) | LEDD (mean ± SD) |
|---|---|---|
| Year 1 | HC (n=175) | NA |
| | PDnP (n=433) | 410.333 ± 340.899 |
| | PDP (n=140) | 442.282 ± 313.698 |
| | PDP at baseline (n=32) | 634.3 ± 373.590 |
| Year 2 | HC (n=165) | NA |
| | PDnP (n=405) | 459.074 ± 381.727 |
| | PDP (n=124) | 518.239 ± 317.295 |
| | PDP at baseline (n=25) | 839.105 ± 669.452 |
| Year 3 | HC (n=157) | NA |
| | PDnP (n=378) | 515.733 ± 445.547 |
| | PDP (n=121) | 564.601 ± 324.676 |
| | PDP at baseline (n=27) | 881.481 ± 641.958 |
| Year 4 | HC (n=151) | NA |
| | PDnP (n=353) | 580.590 ± 462.936 |
| | PDP (n=114) | 608.616 ± 283.115 |
| | PDP at baseline (n=21) | 880.567 ± 407.638 |
| Year 5 | HC (n=145) | NA |
| | PDnP (n=316) | 691.922 ± 775.629 |
| | PDP (n=104) | 682.463 ± 292.151 |
| | PDP at baseline (n=20) | 788.075 ± 433.604 |

**eFigure1**. Predicted values of Levodopa equivalent daily dose (LEDD) in mg/day for each patients' group (i.e., PDP = PD with psychosis, PDnP = PD without psychosis, PDPbaseline = PD with psychosis at baseline) per study year from year 1 to year 5.

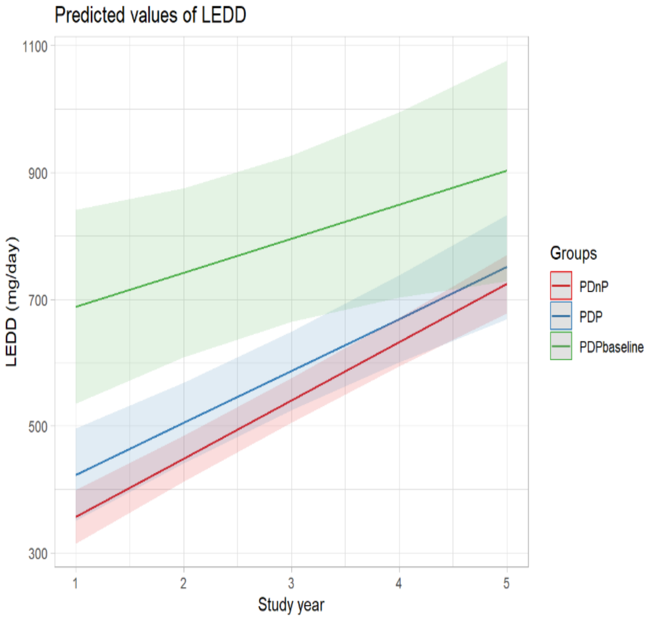

#### *Antipsychotics in PD patients*

The most commonly used antipsychotics were quetiapine, clozapine and olanzapine in the present sample. Both PD and PD psychosis patients reported to take these medications for different reasons. These data are available upon request to the co-authors. eTable2 reports the number of patients in each group who took antipsychotics.

**eTable2.** Table reports the number and percentage (%) of patients on antipsychotics, throughout the study time (from baseline to year 5).

| Group | Quetiapine | Clozapine | Olanzapine |
|---|---|---|---|
| HC (n=2) | 2 (100%) | | |
| PDnP (n=20) | 15 (75%) | 4 (20%) | 1 (5%) |
| PDP (n=18) | 18 (100%) | | |
| PDP at baseline (n=10) | 7 (70%) | 3 (30%) | |

### Supplementary Material 2

#### *Differences between PD psychosis patients at baseline*

Differences between PD psychosis patients at baseline, PDP (i.e., PD patients who developed psychosis after baseline visit) and PD patients without psychosis are reported in eTable3.

**eTable3**. Sample characteristics for PDnP, PDP and PDP at baseline patients. Means and standard deviations (SDs) are reported, unless otherwise specified.

| | **HC (n=187)** | **PDnP (n=503)** | **PDP (n=140)** | **PDP at baseline (n=33)** | **Significance test [a] [*]** | **PDnP vs PDP** | **PDnP vs PDP baseline** | **PDP vs PDP baseline** |
|---|---|---|---|---|---|---|---|---|
| Age (years) (mean ± SD) | 60.324 ± 11.071 | 61.513 ± 10.513 | 62.104 ± 9.449 | 57.854 ± 11.408 | F(3) = 2.042, *P*=0.106 | - | - | - |
| Gender (male, n, %) | 119 (63.6%) | 292 (58.1%) | 82 (58.6%) | 21 (63.6%) | Pearson's Chi-squared test $\chi 2(3) = 2.049$, *P*=0.562 | - | - | - |
| Ethnicity | | | | | Pearson's Chi-squared test $\chi 2(3) = 20.038$, *P*=0.018 | - | - | - |
| *White (n, %)* | 172 (91.9%) | 478 (95%) | 131 (93.6%) | 31 (93.9%) | | | | |
| *Black (n, %)* | 10 (5.3%) | 3 (0.6%) | 4 (2.9%) | 0 | | | | |
| *Asian (n, %)* | 1 (0.5%) | 6 (1.2%) | 2 (1.4%) | 0 | | | | |
| *Other (n, %)* | 4 (2.1%) | 16 (3.2%) | 3 (2.1%) | 2 (6.0%) | | | | |
| PD onset (age in years) (mean ± SD) | - | 58.771 ± 10.769 | 58.968 ± 9.899 | 52.554 ± .389 | F(2)= 5.103, *P*=0.006 | - | *P*=0.005 | *P* =0.008 |
| PD diagnosis (age in years) (mean ± SD) | - | 60.239 ± 10.540 | 60.579 ± 9.641 | 54.979 ± 11.459 | F(2)= 4.181, *P*=0.016 | - | *P* =0.015 | *P* =0.017 |
| Years of education (mean ± SD) | 16.059 ± 2.867 | 15.505 ± 3.398 | 15.057 ± 3.834 | 16.606 ± 4.423 | F(3) = 3.396, *P* = 0.018 | *P* =0.561 | *P* =0.254 | *P* =0.073 |
| PD duration (months) (mean ± SD) | - | 17.271 ± 21.840 | 21.094 ± 24.424 | 38.220 ± 28.284 | F(2) = 16.261, *P*=0.002 | *P* =0.229 | *P* =0.002 | *P* =0.002 |
| MoCA (mean ± SD) | 28.230 ± 1.120<br>28 (26-30) | 26.801 ± 2.625<br>27 (13-30) | 26.964 ± 2.757<br>28 (16-30) | 27.000 ± 2.733<br>27 (21-30) | $\chi 2(3) = 41.829$, *P* < 0.001 | *P* =0.439 | *P* =0.695 | *P* =0.896 |
| HVLT immediate recall (mean ± SD) | 26.075 ± 4.503 | 24.574 ± 4.915 | 24.036 ± 5.349 | 23.727 ± 6.326 | F(3) = 6.046, *P* < 0.001 | *P* =0.9 | *P* =0.9 | *P* =0.9 |

| | | | | | | | | |
|---|---|---|---|---|---|---|---|---|
| HVLT delayed recall (mean ± SD) | 9.312 ± 2.299 | 8.340 ± 2.713 | 8.143 ± 2.799 | 8.697 ± 2.710 | χ2(3) = 21.876, *P* < 0.001 | *P* =0.509 | *P* =0.464 | *P* =0.387 |
| HVLT discrimination (mean ± SD) | 10.048 ± 2.921<br>10 (2-12) | 9.654 ± 2.873<br>9 (0-12) | 9.536 ± 2.563<br>8 (0-12) | 9.636 ± 2.956<br>9 (2-12) | F(3) =1.129, *P* = 0.336 | - | - | - |
| HVLT recognition (mean ± SD) | 11.489 ± 0.833<br>12 (8-12) | 11.167 ± 1.360<br>12 (0-12) | 11.086 ± 1.427<br>12 (4-12) | 11.121 ± 1.409<br>12 (5-12) | χ2(3) = 9.069, *P* = 0.028 | *P* =0.954 | *P* =0.954 | *P* =0.954 |
| Letter number sequence (mean ± SD) | 10.925 ± 2.563 | 10.447 ± 2.780 | 9.850 ± 2.650 | 10.576 ± 3.298 | F(3) = 4.138, *P* = 0.006 | *P* =0.138 | *P* =0.900 | *P* =0.900 |
| Benton judgement of line orientation (BJLOT) (mean ± SD) | 13.134 ± 1.983<br>14 (4-15) | 12.431 ± 2.501<br>13 (0-15) | 12.150 ± 2.603<br>13 (4-15) | 11.818 ± 2.604<br>12 (6-15) | χ2(3) = 18.153, *P* < 0.001 | *P* =0.334 | *P* =0.226 | *P* =0.441 |
| Symbol digit modality (SDM) (mean ± SD) | 47.199 ± 10.493 | 40.896 ± 9.990 | 39.557 ± 11.319 | 39.879 ± 14.124 | F(3) = 19.94, *P* < 0.001 | *P* =0.900 | *P* =0.900 | *P* =0.900 |
| Semantic fluency test (total) (mean ± SD) | 52.280 ± 11.015 | 49.110 ± 11.559 | 49.864 ± 13.225 | 45.758 ± 11.649 | F(3) = 4.636, *P* = 0.003 | *P* =0.900 | *P* =0.676 | *P* =0.423 |
| Depression (GDS) (mean ± SD) | 1.294 ± 2.126<br>1 (0-15) | 2.586 ± 2.738<br>2 (0-14) | 3.081 ± 3.082<br>2 (0-12) | 3.438 ± 3.162<br>3 (0-13) | χ2(3) = 68.275, *P* < 0.001 | *P* =0.14 | *P* =0.12 | *P* =0.44 |
| Sleep (ESS) (mean ± SD) | 5.724 ± 3.448<br>5 (0-19) | 5.640 ± 3.636<br>5 (0-21) | 6.771 ± 4.091<br>6 (0-19) | 9.667 ± 5.109<br>9 (3-21) | χ2(3) = 27.111, *P*<0.001 | *P* =0.008 | *P* <0.001 | *P* =0.006 |
| REM behaviour (mean ± SD) | 2.823 ± 2.253<br>2 (0-11) | 3.931 ± 2.765<br>3 (0-13) | 5.071 ± 2.868<br>5 (0-14) | 6.061 ± 3.482<br>6 (1-14) | χ2(3) = 69.394, *P* < 0.001 | *P* <0.001 | *P* <0.001 | *P* =0.199 |
| Anxiety (STAI) (mean ± SD) | 57.091 ± 14.216<br>53 (40-105) | 65.873 ± 18.385<br>62 (25-123) | 70.243 ± 20.291<br>67 (40-137) | 80.273 ± 19.480<br>74 (47-121) | χ2(3) = 66.684, *P* < 0.001, | *P* =0.032 | *P* <0.001 | *P* =0.009 |
| UPDRS part I scores (mean ± SD) | 2.870 ± 2.948<br>2 (0-17) | 3.739 ± 3.717<br>3 (0-24) | 4.857 ± 4.216<br>4.5 (0-23) | 5.394 ± 3.944<br>4 (1-18) | χ2(3) = 30.458, *P* < 0.001 | *P* =0.004 | *P* =0.005 | *P* =0.343 |
| UPDRS part II scores (mean ± SD) | 0.411 ± 0.929<br>0 (0-5) | 5.723 ± 4.239<br>5 (0-22) | 7.529 ± 4.691<br>6 (1-32_ | 11.091 ± 8.300<br>8 (0-40) | χ2(3) = 391.15, *P* <0.001 | *P* <0.001 | *P* <0.001 | *P* =0.027 |
| UPDRS part III scores (mean ± SD) | 1.043 ± 1.899<br>0 (0-10) | 19.813 ± 9.193<br>19 (0-51) | 21.829 ± 9.986<br>20 (3-48) | 20.636 ± 11.174<br>17 (7-45) | χ2(3) = 427.18, *P* < 0.001 | *P* =0.081 | *P* =0.941 | *P* =0.382 |

| | | | | | | | | |
|---|---|---|---|---|---|---|---|---|
| Rigidity (mean ± SD) | 0.189 ± 0.582<br>0 (0-4) | 3.507 ± 2.568<br>3 (0-11) | 4.129 ± 2.945<br>3 (0-13) | 3.393 ± 3.061<br>3 (0-10) | $\chi2(3) = 335.21$, $P < 0.001$ | $P$ =0.048 | $P$ =0.503 | $P$ =0.175 |
| SCOPA-autonomic (mean ± SD) | 5.859 ± 3.727<br>5 (0-20) | 9.565 ± 6.545<br>8 (0-40) | 13.457 ± 7.438<br>12 (0-38) | 14.636 ± 8.503<br>14 (2-38) | $\chi2(3) = 130.5$, $P < 0.001$ | $P$ <0.001 | $P$ <0.001 | $P$ =0.513 |
| Tremor (mean ± SD) | 0.243 ± 0.794<br>0 (0-7) | 3.922 ± 3.301<br>4 (0-18) | 4.037 ± 3.805<br>3 (0-18) | 2.313 ± 2.507<br>2 (0-9) | $\chi2(3) = 247.47$, $P < 0.001$ | $P$ =0.826 | $P$ =0.008 | $P$ =0.021 |

[a] non-parametric test equivalent for ANOVA, e.g. Kruskal-Wallis test (χ2)
*Where non-parametric tests were used, median and range were also reported.

BJLOT: Benton Judgement Line Orientation test; ESS: Epworth Sleep Scale; GDS: Geriatric Depression Scale; HVLT-R: Hopkins Verbal Learning Test – Revised; MoCA: Montreal Cognitive Assessment; REM: rapid-eye movement; SDM: Symbol Digit Modality test; STAI: State-Trait Anxiety Inventory; SCOPA-autonomic: Scales for Outcomes in Parkinson’s Disease - Autonomic Dysfunction; UPDRS: Unified Parkinson’s Disease Rating Scale (part I-III)

**eTable4.** Number of individuals per group per study year.

| | Baseline | Year 1 | Year 2 | Year 3 | Year 4 | Year 5 |
|---|---|---|---|---|---|---|
| Healthy controls (HC) | 187 | 175 | 165 | 157 | 151 | 145 |
| PD patients without psychosis (PDnP) | 503 | 433 | 405 | 378 | 353 | 316 |
| PD patients with psychosis (PDP) | 140 | 140 | 124 | 121 | 114 | 104 |
| PD patients with psychosis from baseline (PDP at baseline) | 33 | 32 | 25 | 27 | 21 | 20 |

### Supplementary Material 3

*Mixed-effect linear model adjusted for covariates of interest*

Mixed-effect linear model reporting the main effect of group (with HC as reference group, i.e., HC vs PDnP, and HC vs PDP) and the interaction group * time (with HC as reference group) from the analysis adjusted for socio-demographics (age, sex, ethnicity, years of education), depression, sleeping issues (i.e., sleepiness and RBD), and motor symptoms severity. Highlighted in grey the significant results.

**eTable5.** A) HVLT-R tests and B) all other cognitive measures.

A

| | **HVLT-R immediate recall** | | | **HVLT-R delayed recall** | | | **HVLT-R recognition** | | | **HVLT-R discrimination** | | |
|---|---|---|---|---|---|---|---|---|---|---|---|---|
| | **Estimate** | **95% CI** | **P value** | **Estimate** | **95% CI** | **P value** | **Estimate** | **95% CI** | **P value** | **Estimate** | **95% CI** | **P value** |
| **Main effect of group** | | | | | | | | | | | | |
| PDnP vs HC | -0.8 | -1.6, 0.01 | 0.054 | -0.38 | -0.82, 0.05 | 0.083 | -0.12 | -0.32, 0.09 | 0.3 | -0.34 | -0.74, 0.07 | 0.11 |
| PDP vs HC | -0.96 | -2.0, 0.09 | 0.073 | -0.31 | -0.87, 0.24 | 0.3 | -0.11 | -0.37, 0.16 | 0.4 | -0.44 | -0.96, 0.08 | 0.1 |
| **Effect of covariates** | | | | | | | | | | | | |
| Age | -1.5 | -1.8, -1.2 | <0.001 | -0.8 | -0.95, -0.65 | <0.001 | -0.27 | -0.33, -0.20 | <0.001 | -0.39 | -0.52, -0.27 | <0.001 |
| Sex | 0.25 | 0.06, 0.45 | 0.01 | 0.19 | 0.09, 0.29 | <0.001 | 0.08 | 0.02, 0.13 | 0.006 | 0.26 | 0.15, 0.37 | <0.001 |
| Ethnicity | -0.28 | -0.56, 0.00 | 0.05 | -0.16 | -0.31, -0.01 | 0.038 | -0.04 | -0.10, 0.01 | 0.14 | -0.07 | -0.19, 0.04 | 0.2 |
| Education (years) | 1 | 0.74, 1.3 | <0.001 | 0.54 | 0.39, 0.69 | <0.001 | 0.19 | 0.13, 0.25 | <0.001 | 0.16 | 0.03, 0.28 | 0.012 |
| Depression | -0.24 | -0.41, -0.07 | 0.007 | -0.2 | -0.29, -0.11 | <0.001 | -0.05 | -0.10, 0.01 | 0.081 | -0.13 | -0.23, -0.02 | 0.015 |
| RBD | -0.12 | -0.31, 0.08 | 0.2 | -0.02 | -0.13, 0.08 | 0.6 | -0.03 | -0.09, 0.03 | 0.3 | -0.07 | -0.18, 0.04 | 0.2 |
| ESS | -0.02 | -0.20, 0.17 | 0.8 | -0.01 | -0.11, 0.09 | 0.8 | -0.02 | -0.07, 0.03 | 0.5 | 0.02 | -0.08, 0.13 | 0.6 |
| UPDRS part 3 | -0.24 | -0.45, -0.02 | 0.029 | -0.14 | -0.25, -0.03 | 0.014 | -0.04 | -0.11, 0.02 | 0.2 | 0.25 | 0.12, 0.37 | <0.001 |
| **Interaction group * time (Study years)** | | | | | | | | | | | | |
| PDnP * YEAR | -0.13 | -0.29, 0.03 | 0.1 | -0.03 | -0.11, 0.06 | 0.5 | -0.02 | -0.07, 0.04 | 0.5 | -0.19 | -0.29, -0.08 | <0.001 |
| PDP * YEAR | -0.42 | -0.63, -0.21 | <0.001 | -0.17 | -0.28, -0.06 | 0.002 | -0.03 | -0.10, 0.05 | 0.5 | -0.23 | -0.37, -0.10 | <0.001 |

B

| | BJLOT | | | Semantic Fluency | | | SDM | | | LNS | | | MoCA | | |
|---|---|---|---|---|---|---|---|---|---|---|---|---|---|---|---|
| | **Estimate** | **95% CI** | **P value** | **Estimate** | **95% CI** | **P value** | **Estimate** | **95% CI** | **P value** | **Estimate** | **95% CI** | **P value** | **Estimate** | **95% CI** | **P value** |
| **Main effect of group** | | | | | | | | | | | | | | | |
| PDnP vs HC | -0.12 | -0.64, 0.41 | 0.7 | -0.74 | -2.8, 1.3 | 0.5 | -3.8 | -5.4, -2.2 | <0.001 | 0.04 | -0.47, 0.54 | 0.9 | -0.58 | -1.0, -0.14 | 0.009 |
| PDP vs HC | -0.16 | -0.83, 0.51 | 0.6 | 0.67 | -1.9, 3.2 | 0.6 | -4.7 | -6.7, -2.7 | <0.001 | -0.21 | -0.86, 0.43 | 0.5 | -0.59 | -1.1, -0.04 | 0.037 |
| **Effect of covariates** | | | | | | | | | | | | | | | |
| Age | -0.39 | -0.56, -0.22 | <0.001 | -2.8 | -3.5, -2.1 | <0.001 | -4.3 | -4.9, -3.8 | <0.001 | -0.87 | -1.0, -0.70 | <0.001 | -0.78 | -0.93, -0.62 | <0.001 |
| Sex | 0.25 | 0.12, 0.39 | <0.001 | 2 | 1.6, 2.5 | <0.001 | 0.47 | 0.14, 0.81 | 0.006 | 0.47 | 0.34, 0.60 | <0.001 | 0.19 | 0.09, 0.29 | <0.001 |
| Ethnicity | -0.09 | -0.25, 0.08 | 0.3 | -1.1 | -1.8, -0.43 | 0.002 | -0.08 | -0.65, 0.49 | 0.8 | -0.15 | -0.31, 0.01 | 0.074 | -0.2 | -0.35, -0.05 | 0.009 |
| Education (years) | 0.47 | 0.30, 0.64 | <0.001 | 2.5 | 1.8, 3.2 | <0.001 | 2.9 | 2.3, 3.5 | <0.001 | 0.51 | 0.34, 0.68 | <0.001 | 0.39 | 0.24, 0.54 | <0.001 |
| Depression | -0.27 | -0.40, -0.15 | <0.001 | -0.92 | -1.3, -0.51 | <0.001 | -0.75 | -1.0, -0.46 | <0.001 | -0.16 | -0.27, -0.04 | 0.007 | -0.15 | -0.24, -0.07 | <0.001 |
| RBD | -0.13 | -0.26, 0.01 | 0.068 | -0.44 | -0.91, 0.03 | 0.066 | -0.32 | -0.66, 0.01 | 0.06 | -0.15 | -0.28, -0.02 | 0.02 | -0.06 | -0.16, 0.04 | 0.2 |
| Sleepiness (ESS) | 0.08 | -0.05, 0.21 | 0.2 | 0.17 | -0.28, 0.61 | 0.5 | -0.18 | -0.49, 0.14 | 0.3 | 0.04 | -0.08, 0.16 | 0.5 | -0.13 | -0.23, -0.04 | 0.007 |
| UPDRS part 3 | 0.12 | -0.04, 0.27 | 0.14 | -0.34 | -0.85, 0.17 | 0.2 | -1 | -1.4, -0.65 | <0.001 | 0.04 | -0.10, 0.19 | 0.5 | -0.23 | -0.34, -0.12 | <0.001 |
| **Interaction group * time (Study years)** | | | | | | | | | | | | | | | |
| PDnP * YEAR | -0.41 | -0.54, -0.29 | <0.001 | -1.1 | -1.4, -0.67 | <0.001 | -0.17 | -0.43, 0.09 | 0.2 | -0.44 | -0.55, -0.33 | <0.001 | 0.15 | 0.07, 0.23 | <0.001 |
| PDP * YEAR | -0.59 | -0.75, -0.44 | <0.001 | -1.8 | -2.3, -1.3 | <0.001 | -0.51 | -0.85, -0.17 | 0.003 | -0.57 | -0.71, -0.42 | <0.001 | -0.06 | -0.16, 0.05 | 0.3 |

BJLOT: Benton Judgement Line Orientation test; ESS: Epworth Sleep Scale; GDS: Geriatric Depression Scale; HVLT-R: Hopkins Verbal Learning Test – Revised; MoCA: Montreal Cognitive Assessment; REM: rapid-eye movement; SDM: Symbol Digit Modality test; UPDRS: Unified Parkinson's Disease Rating Scale (part I-III)
